# Systematic differences in TB treatment outcomes across in Brazil by patient- and area-related factors: an analysis of national disease registry data

**DOI:** 10.1101/2024.11.26.24317982

**Authors:** Do Kyung Ryuk, Daniele M Pelissari, Kleydson Alves, Luiza Ohana Harada, Patricia Bartholomay Oliveira, Fernanda D C Johansen, Ethel L N Maciel, Marcia C Castro, Ted Cohen, Mauro Sanchez, Nicolas A Menzies

## Abstract

**Background:** A substantial fraction of individuals initiating tuberculosis (TB) treatment do not successfully complete the regimen. Understanding variation in TB treatment outcomes could reveal opportunities to improve the effectiveness of TB treatment services.

**Methods:** We extracted data on TB treatment outcomes, patient covariates, and location of residence from Brazil’s National Disease Notification Information System, for all new TB patients diagnosed during 2015-2018. We analyzed whether or not patients experienced an unsuccessful treatment outcome (any death on treatment, loss to follow-up, or treatment failure). We constructed a statistical model predicting treatment outcome as a function of patient-level covariates, including socio-demographic factors, co-prevalent health conditions, health behaviors, membership of vulnerable populations, and diagnosed form of TB disease. We used this model to decompose state- and municipality-level variation in treatment outcomes into differences attributable to patient-level factors and area-level factors, respectively.

**Results:** Treatment outcomes data for 259,449 individuals were used for the analysis. Across Brazilian states, variation in unsuccessful treatment due to patient-level factors was substantially less that variation due to area-level factors, with the difference between best and worst performing states 7.1 and 13.3 percentage points for patient-level and area-level factors, respectively. Similar results were estimated at the municipality-level, with 9.3 percentage points separating best and worst performing municipalities according to patient-level factors, and 20.5 percentage points separating best and worst performing municipalities according to area-level factors. Results were similar when we analyzed loss to follow-up as an outcome.

**Conclusions:** The results of this analysis revealed substantial variation in TB treatment outcomes across states and municipalities in Brazil, which could not be explained by differences in patient-level factors. This area-level variation likely reflects the consequences of differences in health system organization, clinical practices, and other socio-environmental factors not reflected in patient-level data. Further research to reveal the reasons for these differences is urgently needed to identify effective approaches to TB care, reduce geographic disparities in treatment effectiveness across Brazil, and increase the fraction of patients who successfully complete TB treatment.

## Background

Tuberculosis (TB) is one of the leading causes of infectious disease deaths, killing over 1 million individuals each year [1]. The majority of these deaths occur among individuals who fail to receive treatment. Even for individuals who start TB treatment, a substantial fraction die or discontinue treatment before completing the regimen, or are recorded as having evidence of treatment failure. Of the 7.3 million individuals diagnosed in 2022 for whom TB treatment data were reported to WHO, 12% experienced an unsuccessful treatment outcome [2]. In addition to the poor health outcomes faced by these patients, loss to follow-up and treatment failure create additional opportunities for onward transmission, as well as the development and propagation of TB drug resistance.

Understanding the factors that determine TB treatment outcomes and addressing barriers to high-quality care are important steps for improving the effectiveness of TB services. Analysis of treatment outcomes and other surveillance data has been promoted to identify gaps in TB care cascades, reveal variation in outcomes, and target programmatic action towards areas with the greatest opportunities for improvement [3, 4]. In a number of high TB burden countries, routinely collected TB outcomes data have been used to quantify sub-national differences in the fraction of patients successfully completing treatment [5-11].

In Brazil, TB epidemiology and health services have been shown to vary across the country, with substantial differences in TB incidence, mortality, and case detection rates reported at state and municipal level [12-17]. Similarly, subnational differences have also been reported for TB treatment outcomes [18-20]. Some part of this variation will be explained by differences in individual-level TB risk determinants, and several studies have reported systematic differences in risks of treatment mortality and loss to follow-up according to individual demographics, health-related behaviors, and co-prevalent medical conditions [20-23]. However, those studies that have simultaneously considered both patient-level and spatial factors have found substantial residual variation even after patient-level factors are considered [19, 20], with one study finding the odds of unsuccessful treatment (controlling for patient-level factors) to vary by a factor of 2.9 between best and worst performing states [20].

While some patient-level differences in treatment outcomes will reflect causal processes that are difficult to address (for example, the elevated TB case fatality associated with advanced age), differences in average treatment outcomes between geographic areas could reflect modifiable features of health service organization or practices, which could be targeted to improve overall programmatic outcomes and reduce location-based disparities. Similarly, areas with above-average outcomes could provide examples of successful practices that could be promoted for broader adoption. In this study, we conducted an analysis to systematically quantify subnational differences in TB treatment outcomes in Brazil, adjusted for the range of patient-level factors found to be associated with treatment outcomes in past studies. Based on this analysis, we decompose overall state- and municipality-level differences in treatment outcomes to describe the components attributable to patient-level and area-level factors, and identify the highest- and lowest-performing locations according to each of these metrics.

## Methods

### Study population and data

The study population represented persons without a prior history of TB who were diagnosed with TB and initiated on treatment in Brazil during the years 2015-2018. We obtained data on this study population from Brazil’s National Disease Notification Information System (SINAN) [24]. For each individual we extracted variables describing features of disease presentation and treatment, demographic and socio-economic factors, the presence of co-morbidities, and health-related behaviors. We assigned patients to municipalities based on their recorded municipality of residence at the date of TB notification. For individuals for whom the residential location was not recorded in SINAN (either missing, or could not be linked to a municipality), we used the municipality where the individual received treatment as a proxy for their residence.

### Definition of unsuccessful treatment outcome

We analyzed a binary outcome indicating whether individuals in the study population experienced an unsuccessful treatment outcome. Individuals were categorized as having an unsuccessful treatment outcome if they were recorded as having died from TB or other cause during TB treatment, experienced regimen failure (positive sputum smear or culture in the 4th month or two consecutive months after the 4th month of treatment initiation), or were lost to follow-up (non-attendance at scheduled treatment visits for 30 or more days after treatment initiation). Individuals recorded as experiencing treatment success represented individuals completing the treatment regimen without evidence of regimen failure. We excluded individuals who could not be assigned to unsuccessful or successful treatment outcome based on these definitions. In secondary analyses, we examined a binary indicator for whether or not the individual was lost to follow-up, using the same study sample as the main analysis.

### Adjustment for patient-level factors

We constructed statistical models predicting the treatment outcome as a function of patient-level factors. These included variables previously associated with differences in TB treatment outcomes in this setting: socio-demographic factors (sex, age group, educational level, self-declared race), co-prevalent health conditions (HIV, diabetes), health behaviors (illicit drug use, alcohol use, smoking), membership of vulnerable populations (incarcerated, homeless, immigrant) and diagnosed form of TB disease (pulmonary, extrapulmonary, or both) [20].

We trained and evaluated a range of prediction models based on linear regression, logistic regression, boosted regression trees, and ridge regression. We assessed out-of-sample predictive performance via 10-fold cross-validation, and compared models in terms of the Brier score, C-statistic, visual inspection of calibration plots, calibration intercept and slope, and Hosmer-Lemeshow goodness-of-fit statistic. We selected the best-fitting model as the one that minimized overall predictive error (assessed via the Brier score). As a final validation step, we re-fit this model to data for the years 2015-2017 and assessed how well this model predicted 2018 treatment outcomes data not used for model fitting. The final predictive model was constructed by fitting the chosen model to all data (2015-2018). With this final model, we estimated the probability of an unsuccessful treatment outcome for each individual in the study cohort.

### Estimation of state-level outcomes

We decomposed inter-state variation in the fraction of patients experiencing an unsuccessful treatment outcome into a component attributable to patient-level factors and a second component attributable to area-level factors. We defined *E_i_^All^* as the excess risk of unsuccessful treatment for state *i* compared to the national mean, *E_i_^Patient^* as the excess risk of unsuccessful treatment attributable to patient-level factors, and *E_i_^Area^* as the excess risk of unsuccessful treatment attributable to area-level factors, with *E_i_^All^* equal to the sum of *E_i_^Patient^* and *E_i_^Area^*. To estimate each of these outcomes we calculated:

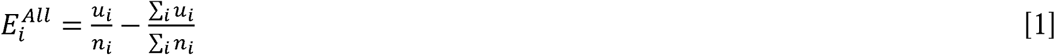

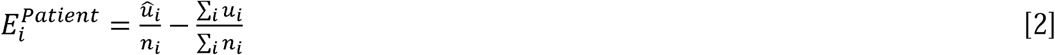

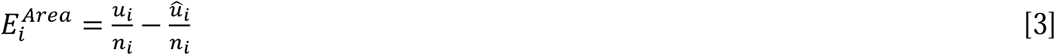

where *u_i_* and *n_i_* represent the observed number of unsuccessful treatment outcomes and total number of observations for state *i* over the study period, respectively, and 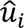 represents the predicted number of unsuccessful treatment outcomes for the same period, calculated by summing the probabilities generated by the predictive model. We multiplied these values by 100 so they represented percentage point differences in the proportion of individuals experiencing unsuccessful treatment outcomes, and used these results to describe the relative contribution of patient-level and area-level factors to overall differences in treatment outcomes across states.

### Estimation of municipality-level outcomes

We modified this approach to estimate *E_i_^All^*, *E_i_^Patient^* and *E_i_^Area^* for municipalities. As some municipalities have small numbers of treated patients, using the approach used for states could result in noisy estimates of *E_i_^Area^*. Instead, we fit a binomial random effects model for the fraction of patients experiencing unsuccessful treatment in each municipality, with the predicted fraction experiencing unsuccessful treatment 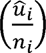 used as an offset term. Based on this specification, the fitted random effect term for each municipality represented the estimated difference in municipality-level outcomes due to area-level factors (*E_i_^Area^*). For municipalities with small numbers of cases the random effect specification pooled this estimate towards the overall mean, while for municipalities with large number of cases this pooling effect was negligible. *E_i_^Patient^* was calculated in the same way as described for states, and *E_i_^All^* was calculated as the sum of *E_i_^Patient^* and *E_i_^Area^*. We used the municipality-level results to describe spatial patterns in *E_i_^Patient^* and *E_i_^Area^* across Brazil.

### Uncertainty analysis and secondary outcomes

We quantified uncertainty in the estimates for each outcome by recalculating results for 10,000 bootstrap samples of the underlying patient data and estimated 95% uncertainty intervals from the 2.5^th^ and 97.5^th^ percentiles of the distribution of bootstrap results for each outcome.

We repeated these analytic steps for an alternative outcome defined as the fraction of TB patients lost to follow-up, with the rationale that this may be the contributor to unsuccessful treatment outcomes that is more amenable to programmatic action.

## Results

There were 356,119 individuals treated for TB over the 2015-2018 study period. We excluded patients previously diagnosed with TB (n=68,519), patients diagnosed with rifampicin resistance (n=2,584), patients who had a change in regimen due to adverse event or identified drug resistance (n=2,019), patients transferred between providers during therapy (n=20,306), patients diagnosed with TB post-mortem (n=2,695), patients with missing treatment outcome values (n=10,786), patients with illogical combinations of exposure variables (n=56), and patients who could not be linked to a municipality (n=42). Records for 259,449 individuals were included in the analysis, representing individuals treated for TB for whom treatment outcomes could be categorized as one of treatment completion, loss to follow-up, death during treatment, or regimen failure. Table 1 reports the distribution of individuals across patient-level exposure variables. Within this study cohort 19.7% of individuals experienced an unsuccessful treatment outcome (death on treatment 7.8%, regimen failure 0.1%, and loss to follow-up 11.9%).

**Table 1.**
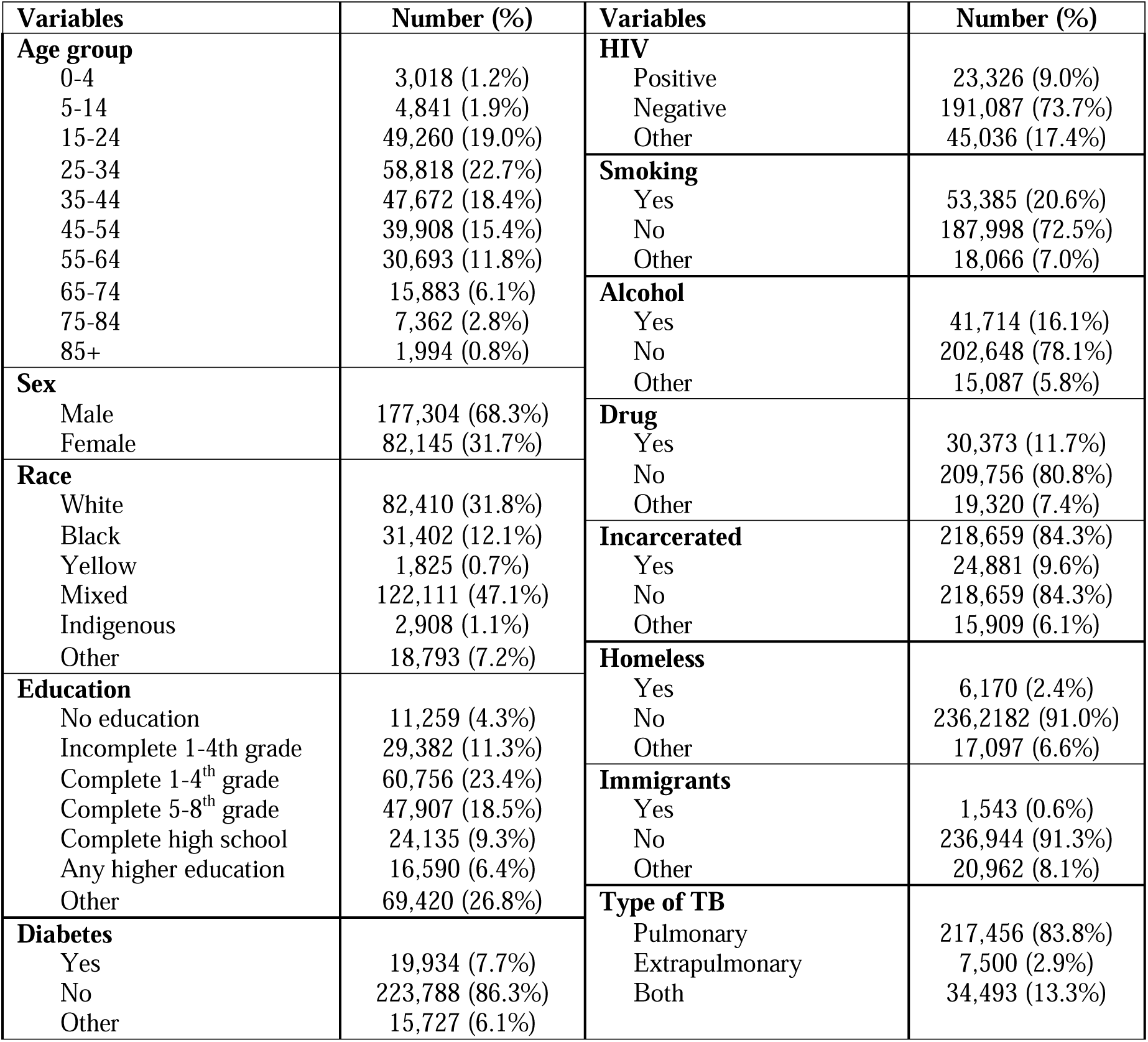
Baseline demographic information of the study population. “Other” category for variables HIV, diabetes, illicit use of drugs, alcohol, smoking, patients in vulnerable circumstances (incarcerated, homeless, immigrant) describe situations where the value was missing or could not otherwise be assigned to Yes or No.

### Prediction model for patient-level factors

We assessed the performance of several candidate prediction models. The best performing model was a logistic regression model with random effects included for all 2-way interactions of patient covariates, with a Brier score of 0.142 (Table 2). This model was among the best-performing for each statistic assessed, and the calibration plot showed good calibration for different levels of predicted risk. When we fit this model to data for 2015-2017 and assessed performance in the 2018 data the out-of-sample predictive accuracy was also good (Figure S1).

**Table 2.**
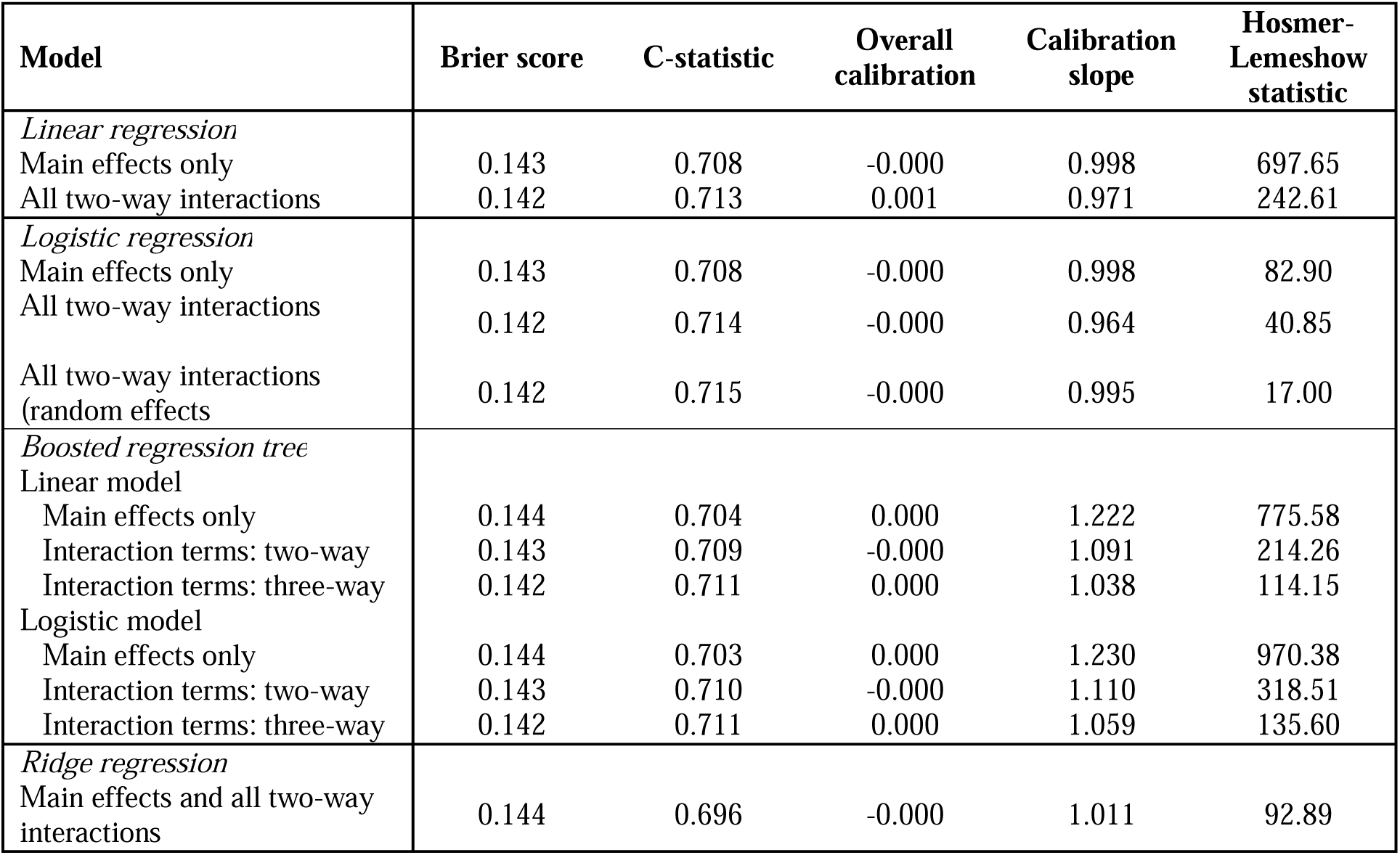
Out-of-sample performance of candidate models used to adjust for patient characteristics. The Brier score quantifies overall predictive performance, with lower values indicating better performance. The C-statistic measures the ability of the model to distinguish patients with different treatment outcomes, with higher values indicating better discrimination. The overall calibration indicates whether the model correctly estimates the overall fraction of the study population experiencing unsuccessful treatment, with a value of zero indicating perfect calibration. When paired with good overall calibration, a calibration slope close to 1 indicates that the model is well calibrated for high- and low-risk individuals. The Hosmer-Lemeshow statistic provides an additional measure of calibration within different risk levels. We estimated this statistic using 10 bins.

| Model | Brier score | C-statistic | Overall calibration | Calibration slope | Hosmer-Lemeshow statistic |
| --- | --- | --- | --- | --- | --- |
| *Linear regression* |  |  |  |  |  |
| Main effects only | 0.143 | 0.708 | -0.000 | 0.998 | 697.65 |
| All two-way interactions | 0.142 | 0.713 | 0.001 | 0.971 | 242.61 |
| *Logistic regression* |  |  |  |  |  |
| Main effects only | 0.143 | 0.708 | -0.000 | 0.998 | 82.90 |
| All two-way interactions | 0.142 | 0.714 | -0.000 | 0.964 | 40.85 |
| All two-way interactions (random effects) | 0.142 | 0.715 | -0.000 | 0.995 | 17.00 |
| *Boosted regression tree* |  |  |  |  |  |
| Linear model |  |  |  |  |  |
| Main effects only | 0.144 | 0.704 | 0.000 | 1.222 | 775.58 |
| Interaction terms: two-way | 0.143 | 0.709 | -0.000 | 1.091 | 214.26 |
| Interaction terms: three-way | 0.142 | 0.711 | 0.000 | 1.038 | 114.15 |
| Logistic model |  |  |  |  |  |
| Main effects only | 0.144 | 0.703 | 0.000 | 1.230 | 970.38 |
| Interaction terms: two-way | 0.143 | 0.710 | -0.000 | 1.110 | 318.51 |
| Interaction terms: three-way | 0.142 | 0.711 | 0.000 | 1.059 | 135.60 |
| *Ridge regression* |  |  |  |  |  |
| Main effects and all two-way interactions | 0.144 | 0.696 | -0.000 | 1.011 | 92.89 |

### State-level outcomes

The excess risk of unsuccessful treatment was calculated by comparing treatment outcomes in each state to the national average. This overall excess risk was then decomposed into the component attributable to patient-level factors and the component attributable to area-level factors (Table 3). Overall, Rio Grande do Sul had the highest proportion of patients experiencing unsuccessful treatment (7.0 (95% confidence interval (CI): 6.4, 7.7) percentage points excess risk compared to the national average), and Acre had the lowest (12.4 (95% CI: 11.1, 13.7) percentage points below the national average). Differences in treatment outcomes attributed to patient-level factors ranged from a 2.8 (95% CI: 2.5, 3.0) percentage points excess risk in Minas Gerais to a 4.3 (95% CI: 3.9, 4.8) percentage point lower risk for Acre. Differences in treatment outcomes attributed to area-level factors ranged from 5.2 (95% CI: 4.6, 5.9) percentage points excess risk in Rio Grande do Sul to an 8.1 (95% CI: 6.8, 9.4) percentage point lower risk for Acre. Across states, the variation in treatment outcomes due to area-level factors (standard deviation 2.71) was greater than for patient-level factors (standard deviation 1.68).

**Table 3.** Excess risk of unsuccessful treatment outcomes by state, decomposed into the contribution from patient-level and area-level factors, 2015-2018.* * *E^All^* represents the percentage point difference in the fraction of individuals experiencing unsuccessful treatment in each state compared to the national average, such that positive values indicate a higher percentage with unsuccessful treatment. *E^Patient^* represents the excess risk of unsuccessful treatment outcomes due to patient-level factors, and *E^Area^* represents the excess risk of unsuccessful treatment outcomes due to area-level factors, such that *E^All^ = E^Patient^*+ *E^Area^*.

| Name | Excess risk of unsuccessful treatment (percentage points) |  |  |
| --- | --- | --- | --- |
| | Total ($E^{All}$) | Due to patient-level factors ($E^{Patient}$) | Due to area-level factors ($E^{Area}$) |
| Rio Grande do Sul | 7.0 (6.4, 7.7) | 1.8 (1.6, 2.1) | 5.2 (4.6, 5.9) |
| Mato Grosso do Sul | 3.8 (2.4, 5.3) | -0.3 (-0.7, 0.2) | 4.1 (2.7, 5.5) |
| Goiás | 3.1 (1.7, 4.5) | 2.3 (1.8, 2.8) | 0.8 (-0.5, 2.1) |
| Rio de Janeiro | 2.7 (2.3, 3.1) | 0.4 (0.2, 0.5) | 2.3 (1.9, 2.7) |
| Amazonas | 2.7 (1.9, 3.4) | -0.1 (-0.3, 0.1) | 2.7 (2.0, 3.5) |
| Mato Grosso | 1.7 (0.4, 2.9) | 0.7 (0.3, 1.1) | 1.0 (-0.2, 2.2) |
| Minas Gerais | 1.1 (0.3, 1.8) | 2.8 (2.5, 3.0) | -1.7 (-2.4, -1.1) |
| Sergipe | 0.7 (-0.8, 2.3) | 0.2 (-0.3, 0.7) | 0.5 (-1.0, 2.1) |
| Ceará | 0.7 (0.0, 1.4) | 1.0 (0.8, 1.3) | -0.4 (-1.0, 0.3) |
| Pernambuco | 0.6 (0.0, 1.2) | 1.8 (1.6, 2.0) | -1.2 (-1.8, -0.6) |
| Roraima | 0.6 (-2.5, 3.7) | -1.1 (-2.0, -0.2) | 1.7 (-1.3, 4.7) |
| Paraíba | 0.5 (-0.9, 1.9) | 0.4 (-0.1, 0.8) | 0.1 (-1.2, 1.5) |
| Rondônia | -0.3 (-2.0, 1.3) | -1.9 (-2.4, -1.4) | 1.5 (-0.1, 3.3) |
| Rio Grande do Norte | -0.4 (-1.7, 1.0) | 1.2 (-0.8, 1.6) | -1.6 (-2.8, -0.3) |
| Alagoas | -0.6 (-2.0, 0.8) | 0.9 (0.5, 1.3) | -1.5 (-2.8, 0.2) |
| Maranhão | -0.7 (-1.6, 0.2) | -1.6 (-1.9, -1.3) | 0.9 (0.1, 1.8) |
| Santa Catarina | -0.9 (-1.8, 0.1) | -0.8 (-1.1, -0.5) | -0.1 (-1.0, 0.9) |
| Bahia | -1.3 (-1.9, -0.6) | 1.6 (1.4, 1.8) | -2.9 (-3.5, -2.3) |
| Federal District | -1.5 (-3.8, 0.7) | -0.4 (-1.2, 0.4) | -1.1 (-3.3, 1.0) |
| Pará | -1.9 (-2.6, -1.3) | 0.0 (-0.2, 0.2) | -1.9 (-2.6, -1.3) |
| São Paulo | -2.7 (-3.0, -2.4) | -1.7 (-1.8, -1.6) | -1.0 (-1.3, -0.8) |
| Tocantins | -3.0 (-5.8, 0.0) | -2.0 (-2.9, -1.0) | -1.0 (-3.8, 1.8) |
| Paraná | -3.1 (-3.9, -2.3) | -1.5 (-1.8, -1.3) | -1.6 (-2.4, -0.8) |
| Espirito Santo | -3.2 (-4.3, -2.1) | -1.0 (-1.4, -0.6) | -2.2 (-3.2, -1.1) |
| Amapá | -4.4 (-6.9, -1.8) | -3.2 (-4.0, -2.5) | -1.1 (-3.6, 1.4) |
| Piauí | -5.1 (-6.6, -3.6) | 0.6 (0.1, 1.1) | -5.8 (-7.1, -4.3) |
| Acre | -12.4 (-13.7, -11.1) | -4.3 (-4.8, -3.9) | -8.1 (-9.4, -6.8) |

### Municipality-level outcomes

We were able to estimate outcomes for 4,911 of the 5,572 municipalities in Brazil, with the remaining 661 municipalities (12%) excluded as they did not have any TB case notifications meeting the inclusion criteria during the study period. Figure 1 shows the municipality-level variation in treatment outcomes across Brazil, separated into the excess risk attributable to patient-level factors (*E^Patient^*, Figure 1A), and area-level factors (*E^Area^*, Figure 1B). For the excess risk attributable to patient-level factors, the highest risks were estimated for the municipalities of Goiânia (Goiás state, excess risk = 5.4 (95% CI: 4.3, 6.5) percentage points), Belo Horizonte (Minas Gerais state, excess risk = 5.3 (95% CI: 4.6, 6.0) percentage points); and Belford Roxo (Rio de Janeiro state, excess risk = 5.2 (95% CI: 4.4, 6.0) percentage points). The lowest excess risks due to patient-level factors were estimated for the municipalities of Macapá (Amapá state, excess risk = -4.0 (95% CI: -4.9, -3.1) percentage points), Rio Branco (Acre state, excess risk = -4.0 (95% CI: -4.6, -3.3) percentage points), and São Vicente (São Paulo state, excess risk = -3.5 (95% CI: -4.0, -2.9) percentage points). For the excess risk attributable to area-level factors, the highest excess risks were estimated for the municipalities of Porto Alegre (Rio Grande do Sul state, excess risk = 11.4 (95% CI: 10.3, 12.5) percentage points), Campo Grande (Mato Grosso do Sul state, excess risk = 10.8 (95% CI: 8.6, 13.0) percentage points), and Nova Iguaçu (Rio de Janeiro state, excess risk = 10.0 (95% CI: 8.4, 11.6) percentage points). The lowest excess risks were estimated for the municipalities of Rio Branco (Acre state, excess risk = -9.1 (95% CI: -11.4, -6.8) percentage points), Ribeirão Preto (São Paulo state, excess risk = -6.9 (95% CI: -9.5, -4.4) percentage points), and São José dos Campos (São Paulo state, excess risk = -6.6 (95% CI: -9.3, -3.9) percentage points). Figure 2 reports excess risk outcomes for major municipalities, including the state capitals plus the top 50 municipalities by total numbers of TB cases during the study period.

**Figure 1.**
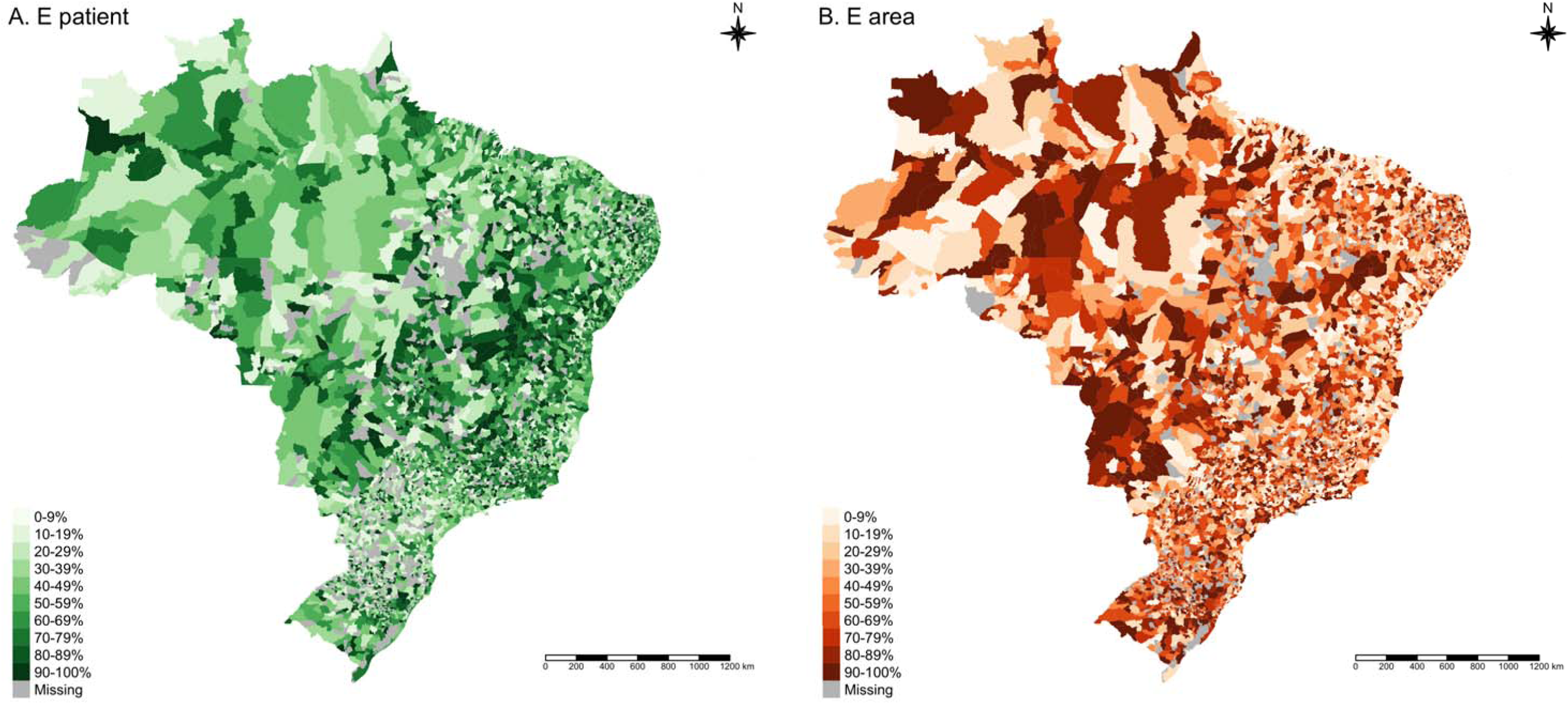
Excess risk of unsuccessful treatment attributable to patient-level and area-level factors, for Brazilian municipalities 2015-2018.* * Panel A shows the distribution of municipalities by decile of excess risk of unsuccessful treatment attributable to patient-level factors (). Panel B shows the distribution of municipalities by decile of excess risk of unsuccessful treatment attributable to area-level factors (). Higher decile (darker shading) indicates a greater risk of unsuccessful treatment

**Figure 2.**
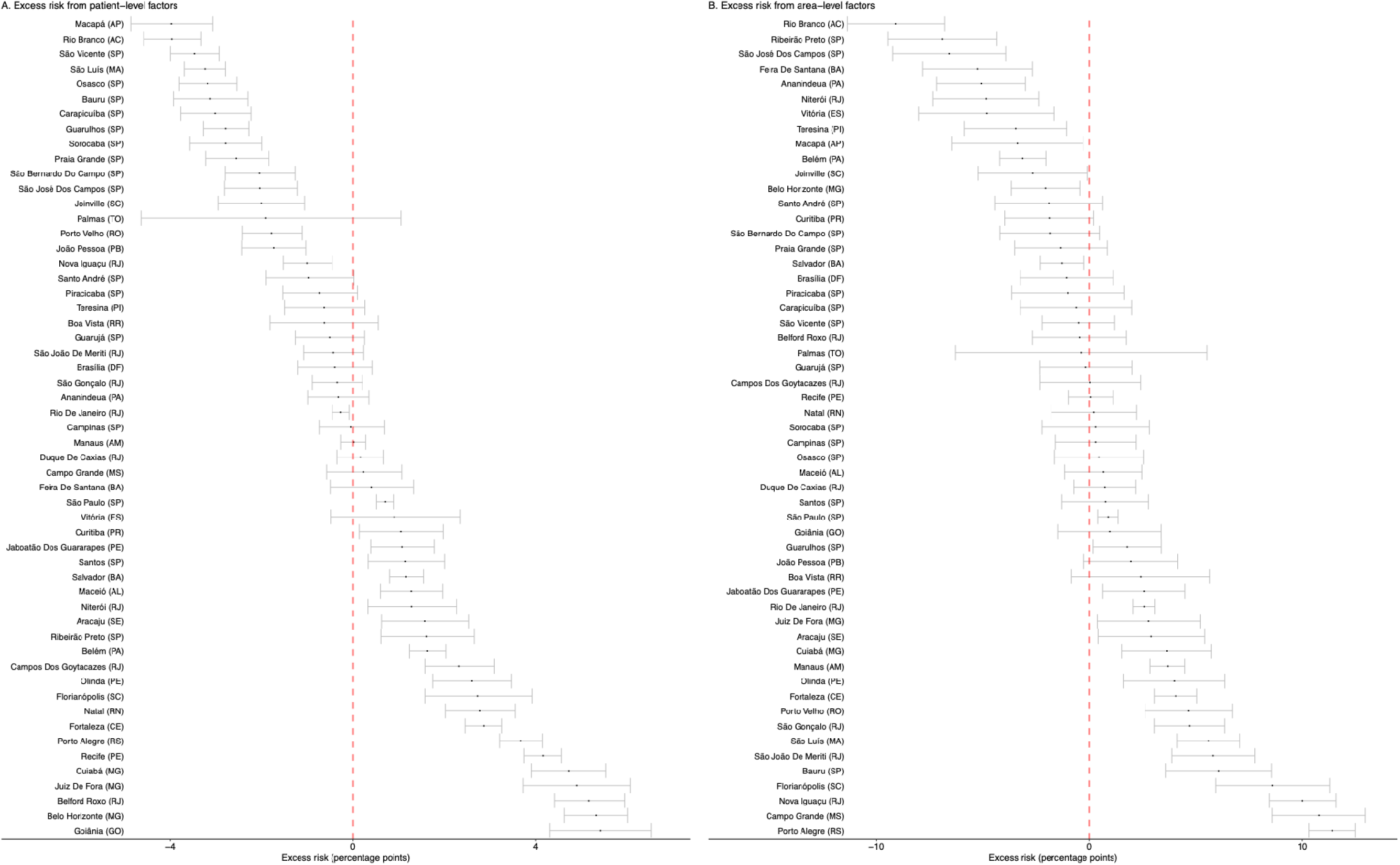
Excess risk of unsuccessful treatment attributable to patient-level and area-level factors, for major municipalities.* * Panel A shows the excess risk of unsuccessful treatment due to patient-level factors (*E^Patient^*). Panel B shows the excess risk of unsuccessful treatment outcomes due to area-level factors (*E^Area^*). Figure shows results for the top 50 municipalities by total TB cases, plus state capitals. Horizontal bars indicate 95% uncertainty intervals. Letters in parentheses indicate the state for each municipality.

Figure S2 shows the joint distribution of municipality-level estimates for excess risk due to patient-level factors and area-level factors, respectively. These two outcomes had a very weak negative correlation (rank correlation coefficient -0.031 (95% CI: -0.059, -0.003), p<0.05), indicating that in general differences in patient-level factors do not explain municipality-level variation in treatment success rates. As with state-level outcomes, the municipal-level variation in treatment outcomes in due to area-level factors (standard deviation 4.27) was greater than for patient-level factors (standard deviation 2.52).

### Secondary outcome: excess risk of loss to follow-up

We re-estimated results for an alternative outcome defined as the excess risk of loss to follow-up. Figure S3 shows the distribution of municipality-level estimates for the excess risk of loss to follow-up attributable to patient-level and area-level factors (Figure S3A and S3B, respectively). Municipality-level results for this secondary outcome were positively associated with the results of the main analysis, with a correlation coefficient of 0.56 (95% CI: 0.55, 0.58); p <0.05) for *E^Patient^* and a correlation coefficient of 0.77 (95% CI: 0.75, 0.78; p<0.05) for *E^Area^*. For *E^All^* the correlation coefficient was 0.64 (95% CI: 0.63, 0.66; p<0.05).

## Discussion

In this study, we assessed how the outcomes of TB treatment vary across Brazil according to patient-level and area-level factors, using data on 259,449 individuals initiating TB treatment over the 2015-2018 period. At both state and municipality level, we found that variation in the fraction of individuals experiencing an unsuccessful treatment outcome due to area-level factors—factors that can’t be explained by differences in patient characteristics—was substantially greater than the variation due to patient-level factors. For example, in our analysis, the difference between best and worst performing states according to patient-level factors represented a 7.1 percentage point increase in the fraction of individuals experiencing an unsuccessful treatment outcome. In contrast, the difference between best and worst performing states according to area-level factors represented a 13.3 percentage point increase, almost twice as great. A similar result was found at the municipality level. This variation due to area-level factors represents a major difference in the numbers of patients experiencing treatment success. For example, if all states were able to improve treatment outcomes to be at least as good as the top quartile of states, this would represent an average 1,142 additional TB patients achieving successful treatment every year in the study population, or 1.8% of total notifications.

By adjusting for differences in treatment outcomes due to patient-level factors, the residual variation (as quantified by *E^Area^*) represents variation that cannot be attributed to differences in the individual risk factors recorded in the notifications data. While some of this variation may reflect the impact of individual risk factors not included in the notification record, it is likely that a major part of this variation represents the impact of differences in health system organization and clinical practices across states and municipalities. Practically, such variation could reflect differences in travel distances and other barriers that patients face to initiate and attend treatment, the range of support services provided during treatment, or the level of effort devoted to following-up on patients who have missed an appointment. It could also reflect differences in treatment access across geographic areas, with delayed treatment initiation as a possible cause of poor treatment outcomes. Beyond healthcare access and quality, area-level variation in treatment outcomes could also include local socio-environmental factors that influence treatment adherence and completion. Such factors may include public initiatives to address social inequalities (e.g., food vouchers, cash transfers), and social protection. Variation in outcomes due to patient-level factors (as quantified by *E^Patient^*) has been investigated in prior studies [20-23], with treatment mortality varying by age and HIV status, and loss to follow-up varying by factors tied to socioeconomic disadvantage (limited education and harmful health behaviors).

In addition to describing the variation in treatment outcomes across Brazil, this analysis identified specific states and municipalities that stood apart from other areas. For patient-level factors, Acre and Amapá states had low excess risks of unsuccessful treatment (>3 percentage points below the national average), while Goiás and Minas Gerais had high excess risks (>2 percentage points above the national average). While it is difficult for TB programs to directly change the risk profile of individuals diagnosed with TB, understanding the differences between these states may reveal important differences in how TB risk is distributed across communities. For area-level factors, Acre and Piauí states had low excess risks of unsuccessful treatment (>5 percentage points below the national average), while Rio Grande do Sul and Mato Grosso do Sul had high excess risks (>4 percentage points above the national average). For the high-performing states, it may be that they have adopted care practices that allow them to achieve better outcomes. If true, these practices could potentially be employed in other areas to improve performance. Less optimistically, it is possible that these states have systematic problems with how treatment outcomes are being reported, of have weaker case detection among the vulnerable populations most at risk of poor treatment outcomes, such that these individuals are never diagnosed and therefore not represented in treatment outcomes data. In both cases, further investigation would be valuable. Similarly, for those states for which this analysis estimated poor outcomes, there may be opportunities to improve treatment outcomes through further investigation to reveal the causes of poor treatment outcomes. A similar approach can be taken to studying municipalities that had results substantially higher or lower than the national average.

While previous studies have examined subnational differences in TB treatment outcomes, this is the first study that we are aware of that has systematically adjusted for differences in treatment outcomes due to patient level factors and decomposed overall rates of unsuccessful treatment to describe the contribution of both patient-level and area-level factors. We were able to do so using by a large and nationally representative dataset, allowing relatively precise inferences about how treatment outcomes vary across Brazil. This analysis also has several limitations. First, while the TB notification data used for this analysis include a range of relevant variables, it is possible that there are other individual-level factors that influence outcomes but that are not recorded as part of the notification. If the prevalence of these factors differs systematically across states and municipalities, their omission from the analysis could bias the decomposition into patient-level and area-level factors. Second, this analysis relies on the validity of treatment outcome reporting, and if there were systematic inconsistencies in how clinicians assign patients to treatment outcome categories, this would undermine the analysis. This is more likely to affect the secondary outcome (loss to follow-up), as a patient who dies while on treatment may be misclassified as lost to follow-up unless there are robust efforts to follow-up on patients who stop attending clinic visits. Similarly, treatment outcomes were unavailable for some patients (treatment outcome recorded as transfer or missing, 9% of total sample) that would otherwise have been included in the analysis. If outcomes for these patients were systematically different from those included in the analysis this could bias the results. Third, we did not attempt to identify the reasons why certain states and municipalities were achieving better or worse outcomes than others. This investigation represents a priority for future work and is a necessary step for translating this study’s findings into programmatic improvements. Fourth, this analysis attributed each patient’s treatment outcomes to their state and municipality of residence. While the approach we used will likely be unproblematic for state-level analysis, it is possible that some municipalities have a non-negligible fraction of patients travelling to neighboring municipalities to receive care.

In summary, our study identified substantial variation in TB treatment outcomes across states and municipalities in Brazil, which could not be explained by differences in patient-level factors. Further research to reveal the reasons for these differences is urgently needed to reduce geographic disparities in TB care across Brazil and increase the fraction of patients who successfully complete TB treatment.

## Conclusion

Using prediction modeling, we adjusted for the case-mix of individuals treated for TB and estimated the variation of TB treatment outcome across state and municipal level in Brazil. We found greater variation in both state and municipal level in the area-level factors compared to the patient-level factors, and this variation is likely a result of differences in how health systems are structured, clinical practices in place and the impact of socio-economic factors. Thus, further research is necessary to understand these differences to shape effective strategies for quality of TB care in regional level and improve TB treatment outcome in Brazil.

## List of Abbreviation

TB: Tuberculosis
WHO: World Health Organization
SINAN: Sistema de Informação de Agravos de Notificação
HIV: Human Immunodeficiency Virus

## Data Availability

All data used in this study is publicly available. The demographic dataset analyzed during the current study are available in IBGE, https://www.ibge.gov.br/. TB case and mortality data can be accessed at the Ministry of Health of Brazil website, https://datasus.saude.gov.br/.

https://datasus.saude.gov.br/

## SUPPLEMENTARY TABLES/FIGURES

**Figure S1.**
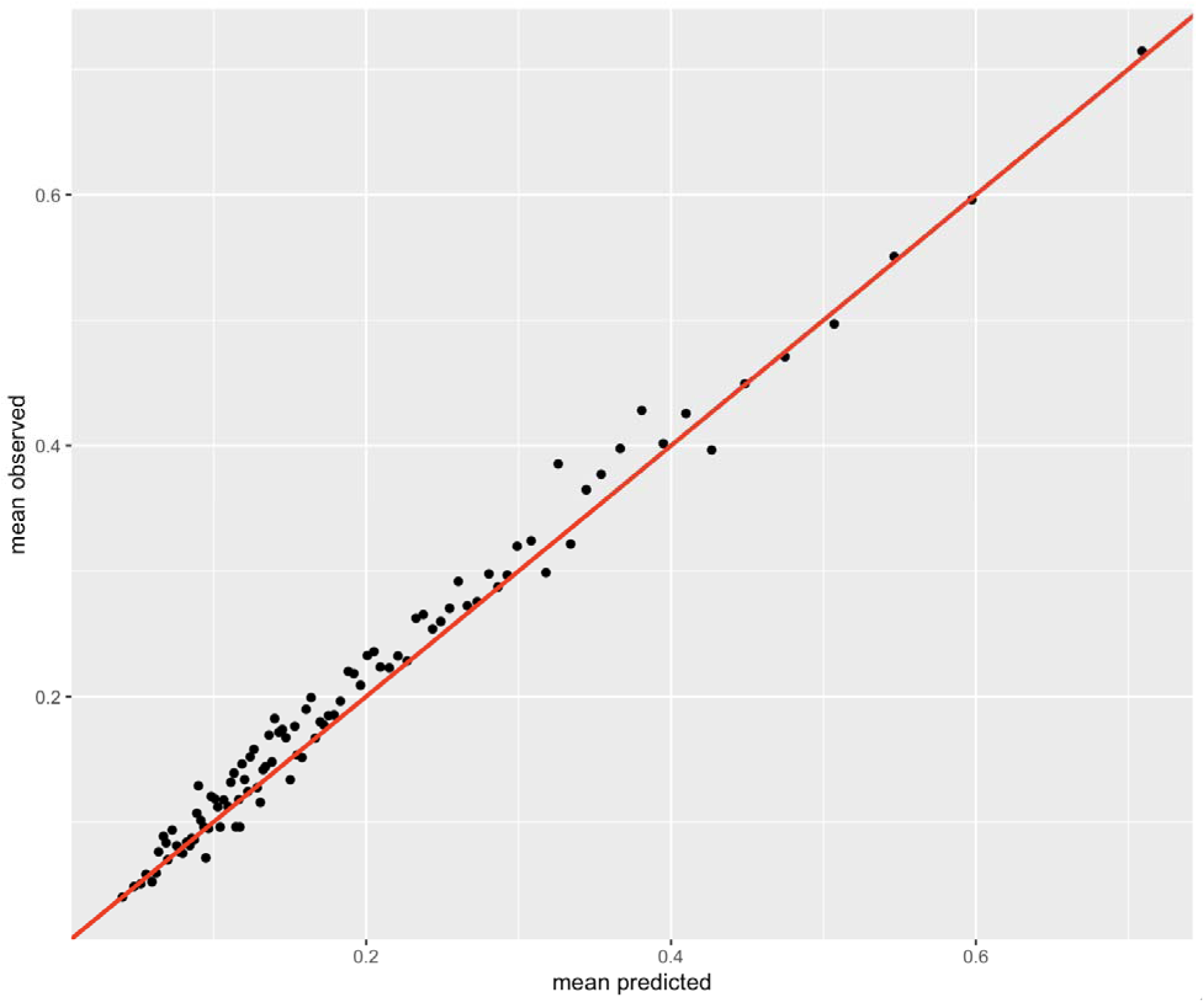
Calibration plot for the selected model.

**Figure S2.**
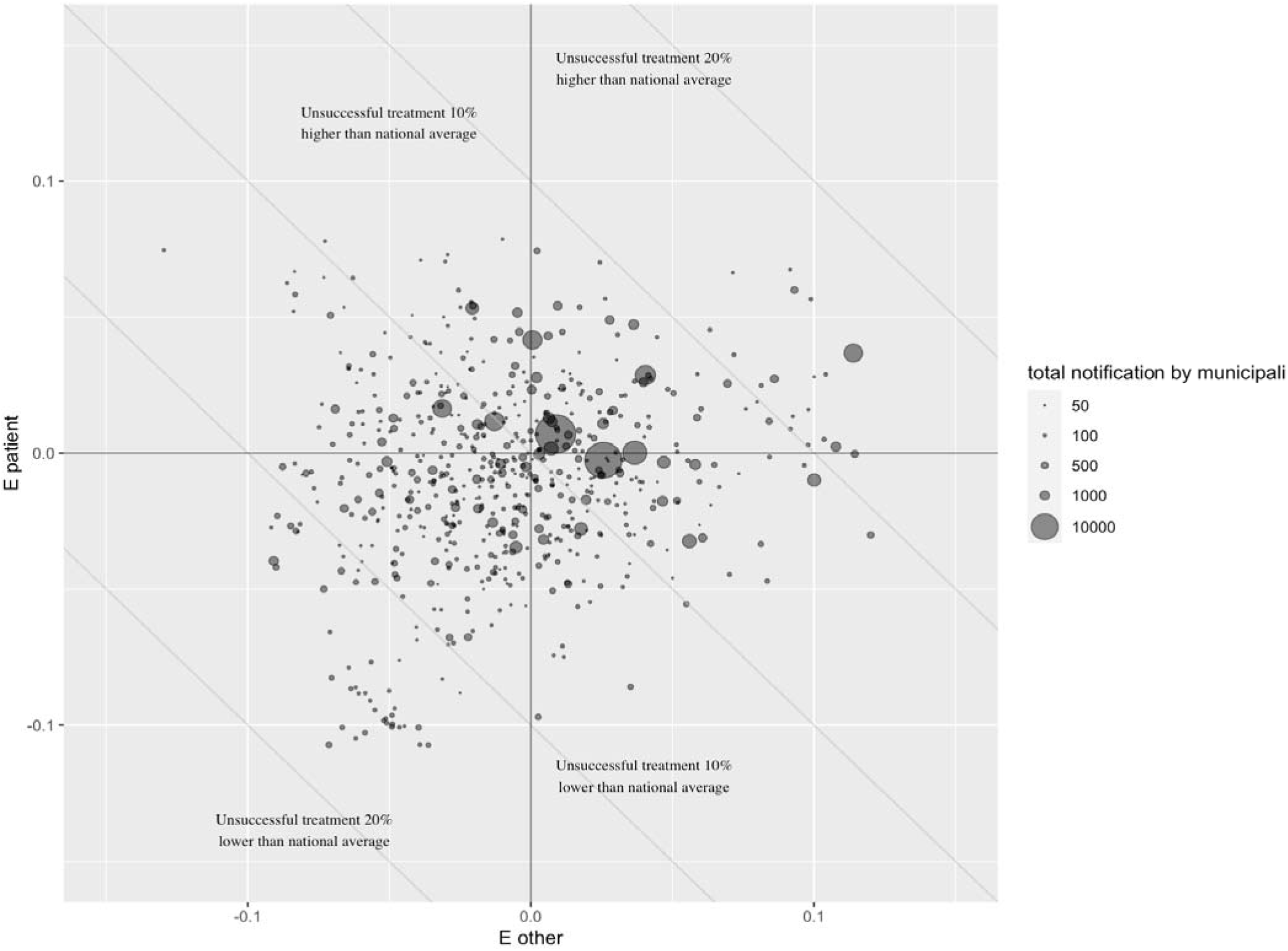
Scatterplot of area-level factors () and patient-level factors () across municipalities.

**Figure S3.**
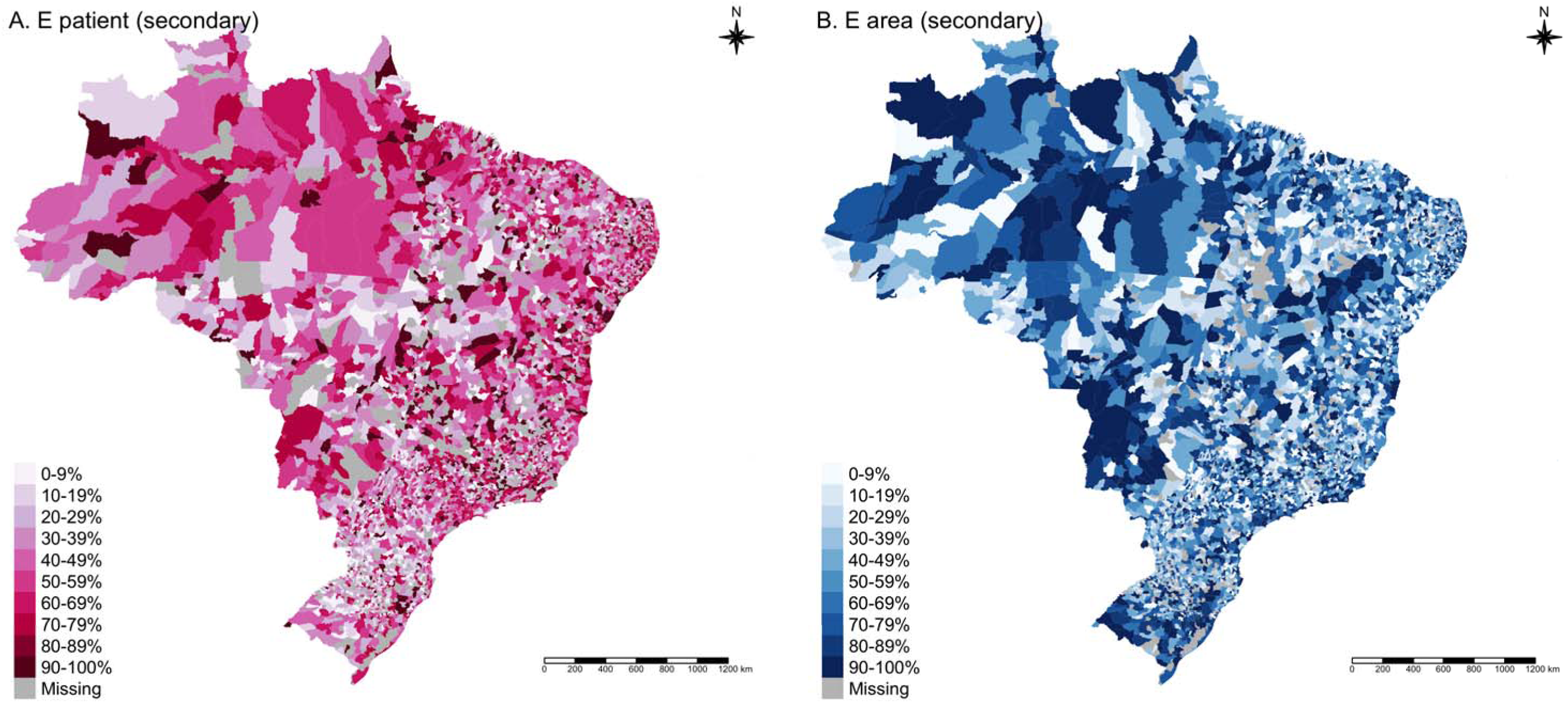
Excess risk of loss to follow-up (secondary outcome) attributable to patient-level and area-level factors, for Brazilian municipalities 2015-2018.* * Panel A shows the distribution of municipalities by decile of excess risk of loss to follow-up attributable to patient-level factors (). Panel B shows the distribution of municipalities by decile of excess risk of loss to follow-up attributable to area-level factors (). Higher decile (darker shading) indicates a greater risk of loss to follow-up.

## References

1. World Health Organization: Global TB Report 2024. In. Geneva; 2024.

2. WHO Global TB Programme: WHO Global TB Database [http://www.who.int/tb/country/data/download/en/]. In. Geveva Switzerland: WHO Global TB Programme; 2024.

3. Faust L, Naidoo P, Caceres-Cardenas G, Ugarte-Gil C, Muyoyeta M, Kerkhoff AD, Nagarajan K, Satyanarayana S, Rakotosamimanana N, Grandjean Lapierre S et al: Improving measurement of tuberculosis care cascades to enhance people-centred care. The Lancet Infectious Diseases 2023, 23(12):e547–e557.

4. Sismanidis C, Shete PB, Lienhardt C, Floyd K, Raviglione M: Harnessing the Power of Data to Guide Local Action and End Tuberculosis. The Journal of Infectious Diseases 2017, 216(suppl_7):S669-S672.

5. Iskandar D, Suwantika AA, Pradipta IS, Postma MJ, van Boven JF: Clinical and economic burden of drug-susceptible tuberculosis in Indonesia: national trends 2017–19. The Lancet Global Health 2023, 11(1):e117–e125.

6. Surya A, Setyaningsih B, Suryani Nasution H, Gita Parwati C, Yuzwar YE, Osberg M, Hanson CL, Hymoff A, Mingkwan P, Makayova J: Quality tuberculosis Care in Indonesia: using patient pathway analysis to optimize public–private collaboration. The Journal of infectious diseases 2017, 216(suppl_7):S724-S732.

7. Alene KA, Viney K, Gray DJ, McBryde ES, Wagnew M, Clements AC: Mapping tuberculosis treatment outcomes in Ethiopia. BMC infectious diseases 2019, 19:1–11.

8. Matthew Adebayo A, Olaiya Adeniyi B, Oluwasanu M, Hassan A, Ada Ajuwon G, Chidinma Ogbuji Q, Adewole D, John Osho A, Olukolade R, Alabi Ladipo O et al: Tuberculosis treatment outcomes and associated factors in two states in Nigeria. Tropical Medicine & International Health 2020, 25(10):1261–1270.

9. Schnaubelt E, Charles M, Richard M, Fitter D, Morose W, Cegielski J: Loss to follow-up among patients receiving anti-tuberculosis treatment, Haiti, 2011–2015. Public health action 2018, 8(4):154-161.

10. Hippner P, Sumner T, Houben RM, Cardenas V, Vassall A, Bozzani F, Mudzengi D, Mvusi L, Churchyard G, White RG: Application of provincial data in mathematical modelling to inform sub-national tuberculosis program decision-making in South Africa. PloS one 2019, 14(1):e0209320.

11. Emani S, Alves K, Alves LC, da Silva DA, Oliveira PB, Castro MC, Cohen T, Couto RM, Sanchez M, Menzies NA: Quantifying gaps in the tuberculosis care cascade in Brazil: A mathematical model study using national program data. PLoS Med 2024, 21(3):e1004361.

12. Chitwood MH, Alves LC, Bartholomay P, Couto RM, Sanchez M, Castro MC, Cohen T, Menzies NA: A spatial-mechanistic model to estimate subnational tuberculosis burden with routinely collected data: An application in Brazilian municipalities. PLOS Global Public Health 2022, 2(9):e0000725.

13. Chitwood MH, Pelissari DM, da Silva GDM, Bartholomay P, Rocha MS, Sanchez M, Arakaki-Sanchez D, Glaziou P, Cohen T, Castro MC: Bayesian evidence synthesis to estimate subnational TB incidence: An application in Brazil. Epidemics 2021, 35:100443.

14. Harling G, Castro MC: A spatial analysis of social and economic determinants of tuberculosis in Brazil. Health & place 2014, 25:56–67.

15. Polidoro M, de Oliveira DC: Prevalence and Spatial Autocorrelation of Tuberculosis in Indigenous People in Brazil, 2002-2022. Journal of Racial and Ethnic Health Disparities 2024:1-8.

16. Berra TZ, Ramos ACV, Alves YM, Tavares RBV, Tartaro AF, Nascimento MCd, Moura HSD, Delpino FM, de Almeida Soares D, Silva RVdS et al: Impact of COVID-19 on Tuberculosis Indicators in Brazil: A Time Series and Spatial Analysis Study. Tropical Medicine and Infectious Disease 2022, 7(9):247.

17. Martins-Melo FR, Bezerra JMT, Barbosa DS, Carneiro M, Andrade KB, Ribeiro ALP, Naghavi M, Werneck GL: The burden of tuberculosis and attributable risk factors in Brazil, 1990– 2017: results from the Global Burden of Disease Study 2017. Population Health Metrics 2020, 18:1-17.

18. Tavares RBV, Berra TZ, Alves YM, Popolin MAP, Ramos ACV, Tártaro AF, de Souza CF, Arcêncio RA: Unsuccessful tuberculosis treatment outcomes across Brazil’s geographical landscape before and during the COVID-19 pandemic: are we truly advancing toward the sustainable development/end TB goal? Infectious Diseases of Poverty 2024, 13(1):17.

19. Prado Junior JC, Medronho RdA: Spatial analysis of tuberculosis cure in primary care in Rio de Janeiro, Brazil. BMC Public Health 2021, 21:1–15.

20. Ryuk DK, Pelissari DM, Alves K, Oliveira PB, Castro MC, Cohen T, Sanchez M, Menzies NA: Predictors of unsuccessful tuberculosis treatment outcomes in Brazil: an analysis of 259,484 patient records. BMC Infectious Diseases 2024, 24(1):531.

21. Barreto-Duarte B, Araújo-Pereira M, Nogueira BM, Sobral L, Rodrigues MM, Queiroz AT, Rocha MS, Nascimento V, Souza AB, Cordeiro-Santos M: Tuberculosis burden and determinants of treatment outcomes according to age in Brazil: a nationwide study of 896,314 cases reported between 2010 and 2019. Frontiers in Medicine 2021, 8:706689.

22. Ridolfi F, Peetluk L, Amorim G, Turner M, Figueiredo M, Cordeiro-Santos M, Cavalcante S, Kritski A, Durovni B, Andrade B: Tuberculosis treatment outcomes in Brazil: different predictors for each type of unsuccessful outcome. Clinical Infectious Diseases 2023, 76(3):e930–e937.

23. Chenciner L, Annerstedt KS, Pescarini JM, Wingfield T: Social and health factors associated with unfavourable treatment outcome in adolescents and young adults with tuberculosis in Brazil: a national retrospective cohort study. The Lancet Global Health 2021, 9(10):e1380–e1390.

24. Brasil Ministério da Saúde: Departamento de Informática do Sistema Único de Saúde. Sistema de Informação de Agravos de Notificação (SINAN) [https://datasus.saude.gov.br/transferencia-de-arquivos/]. In.: Brasilia, Brazil; 2024.

